# A Validation Study for the Successful Isolation Policy in China: a Meta-Analysis in COVID-19

**DOI:** 10.1101/2020.04.15.20065102

**Authors:** Jianghu (James) Dong, Yongdao Zhou, Ying Zhang, Douglas Fraz

**Author notes:** Corresponding author. Address: Department of Biostatistics, College of Public Health, University of Nebraska Medical Center, Omaha, Nebraska, USA, 68198-4375.

## Abstract

COVID-19 is quickly spreading around the world and carries along with it a significant threat to public health. This study sought to apply meta-analysis to more precisely and accurately estimate the basic reproduction number (*R*_0_) of COVID-19 in order to evaluate the effectiveness of the isolation policy across countries, and the corresponding public health capability to care for patients. Prior estimates of *R*_0_ have varied widely and range from 1.95 to 6.49. Utilizing meta-analysis techniques we determined a more robust estimation of 3.05 for *R*_0_, which is substantially larger than that provided by the WHO. We also present the infectious rate standardized to per million population, which has proved to be a good index to determine whether the isolation measures in specific countries are effective. Also, this standardized infection rate can be used to determine whether the current infectious severity status is out of range of the national health capacity. Finally, we utilize our robust estimate of *R*_0_ and the standardized infectious rate to illustrate that the early and aggressive isolation measures enforced by the Chinese government were substantially more effective in controlling the negative impact COVID-19 than the more permissive measures enacted early in Italy and the United States.

## 1 Background

December of 2019 saw an unprecedented spike in the number of cases of lower respiratory tract infections in Wuhan, China. Most patients presented with symptoms resembling viral pneumonia which is characterized by fever, dry cough, dyspnea, fatigue, and body aches. The outbreak in Wuhan was found to be due to a novel strain of COVID-19 with high virulence and transmissibility. On January 12th, 2020, the World Health Organization (WHO) named it as COVID-19 **WHO2020**. COVID-19 is the seventh member of the human-infected coronavirus family **Chan2020**. Other notable coronavirus family members include severe acute respiratory syndrome (SARS) coronavirus and Middle East Respiratory Syndrome (MERS). All have been responsible for massive outbreaks within the last 15 years. As with its predecessors, COVID-19 may be transmitted from person-to-person by droplet or contact as well as the fecal-oral route **Wang2020**. Because it is a novel strain, there is no herd immunity which partially accounts for its rapid spread and unfortunately, the strain is highly virulent, with a mortality rate well over 10-fold that of the illness caused by the seasonal H. influenza virus. By Jan 23, 2020, Wuhan city was quarantined and shortly thereafter all of the other provinces in China announced heavy restrictions on travel. However, COVID-19 had already escaped containment and is rapidly spreading across the world.

As of this writing, there are no known anti-viral medications that directly eliminate COVID-19. In addition to supportive care, when available, infected patients otherwise must rely on the immune response to overcome the novel virus. The innate and adaptive immune response may inhibit and then eliminate the virus by selecting and then producing antibodies specific to the virus epitope(s). Because the virus is novel, Human B-cells have no memory of the viral antigen and thus their response is both delayed and diminutive. Thus, previously uninfected humans, especially older individuals, people with poor immunity or on immunosuppression, or with underlying comorbidities are at relatively high risk of mortality. Scientists are actively working on a vaccine that provides the previously uninfected human host with immunity by injecting a non-virulent epitope of the virus so that the body may develop memory B cells that can rapidly and vigorously attack and eliminate the virus with specific anti-viral antibodies. Unfortunately, even optimistic projections for the delivery date of an effective vaccine against COVID-19 would arrive far too late to impede the massive wave of COVID-19 spread.

In the absence of an effective vaccine or anti-viral regimen to eliminate the virus, the best mechanism to prevent rapid transmission and overutilization of available healthcare resources, which may lead to unnecessary death, is the early and aggressive isolation of infected individuals and at-risk populations. Isolation decreases the effective reproductive number by decreasing the transmissibility of the virus. Strategies to limit the spread of COVID-19 have varied drastically by country and even by state and local governments/institutions in the United States (US). The duration and aggressiveness of isolation necessarily depend on the stage of the outbreak for the country in question. For example, China has reached a buffer period by using early and aggressive quarantine measures, Most of Europe is currently at its peak period, and the US remains in a period of exponential case growth which may be due to late, inconsistent, and relatively permissive isolation measures enacted by states in absence of an early unified federal response.

The number of people who are infected during the peak period depends mainly on the efficacy of quarantine in the absence of a vaccine. So the quarantine has been carried out to decrease the effective reproduction number of COVID-19. From the term of epidemic principles, the virus usually has the initial basic transmissibility *R*_0_. *R*_0_ is an important index to determine the epidemic intensity. So many studies have been carried out to estimate it as we describe in detail below. As the efficacy of quarantine increases, the reproductive number decrease. If the declining trend continues with the assumption of no resurges of the epidemic, the reproduction number will drop below one, meaning that each individual will, on average, infect less than one other individual and subsequently, the epidemic is gradually die off at this time point when the effective reproductive number reaches one. Also, the peak of the infection rate can be delayed or reduced after the government intervention by reducing the effective reproductive number *R*_*t*_, and accordingly, it reduces the strain on healthcare systems which are set to run at near-capacity in absence of an epidemic.

Therefore, the above epidemic scenarios motivated us to investigate the effectiveness of the isolation policy across different countries from real data since it is important for public health to identify effective measures of prevention for COVID-19 spread. This study has three purposes. First, since estimates of *R*_0_ range widely (1.95 to 6.47) in the existing literature, we sought to utilize metaanalyses to determine a more robust estimate for *R*_0_. Second, we standardized the infectious rate to per million population as a more conducive comparison of the distribution of COVID-19 and to more readily show how the infectious rate is beyond the maximum of the health system capacity in some countries. Third, we can show how the relative success of China isolation policy to control the effective reproductive number from the statistical model based on real data. To this end, the results can supply some useful guidelines for controlling the rapid spread of COVID-19 in the world. The rest of this article is organized as follows. The proposed model is introduced in Section 2. Section 3 demonstrates the results from the proposed model. Conclusions are given in Section 4.

## 2 Method

We introduce the related statistical models to solve the question in COVID-19 research in this section. Each virus has a basic reproductive index of *R*_0_. The higher the reproduction index, the greater the spread of the virus in the absence of quarantine (government isolation policy). So, we assumed that the number of infected patients is related to the total population, the effective reproduction number index, and the government isolation policy to stop the rapidly transmitted from person to person. Now let *Y*_*t*_ denote the number of infectious patients in a specific country, and the model for the number of the COVID-19 cases is assumed to be dependent on the total population, the reproduction number index, and the government isolation policy as follows:

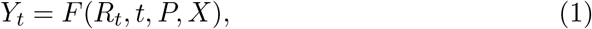

where *R*_*t*_ denotes the effective reproduction index at time *t, P* denotes the total population in the specific country, and *X* denotes the local government such as a balance between the freedom and permissiveness. The function *F* can be a suitable predict function with these variables *P, R*_*t*_, and *X*. For the function *F*, the paper **liu2020** used functional mixed effect model to *Y*_*t*_. We do not focus on the construction of the function *F* in this paper.

The basic reproductive number *R*_0_ is an important pandemic index to indicate infectious intensity, but it is challenging to determine the true value. Many studies have given a wide range of *R*_0_ in the existing literature, and we propose the meta-analysis to estimate it in Section 2.1. Usually, the higher the reproduction number, the more people will be infected given no intervention. We proposed to use the infectious rate per million population from *Y*_*t*_ and *P* to see the pandemic severity in Section 2.2. Also, this standardized infectious rate per million population also allows us to compare the intervention effects of COVID-19 across countries. If the infectious rate exceeds the maximum healthcare capacity, the pandemic will cause a high mortality rate. Therefore, we discuss the relationship between the healthcare capacity and the peak of the infectious rate in Section 3.

### 2.1 The basic reproduction number

As mentioned in Section 1, the basic reproduction number *R*_0_ is an indication of the initial transmissibility probability of a virus. It represents the average number of newly infected patients generated by an already infectious person. For example, if *R*_0_ is 3, then each patient has been spreading the infection to 3 other peoples in theory. We have done a systematic review of the reproduction index, and we find that many studies such as **Wu2019**; **Read2019**; **Imai2020**; **and Riou2020** have been carried out to estimate this important basic reproduction number index from statistical models. Most of these studies were based on the stochastic process and the growth statistical model in the exponential distribution family. The estimated values of *R*_0_ were different from each other with the range was from 1.95 to 6.49. This huge difference of *R*_0_ motivated us to estimate the basic reproduction number by the scientific meta-analysis method, which is a statistical tool that combines the results of multiple scientific studies.

Meta-analysis can be used to address the same question in multiple scientific studies, where each individual study reporting measurement was expected to have some degree of error. So our main aim is to use the meta-analysis approach to derive a pooled estimate closest to the unknown common truth. A benefit of this approach was allowing us to have the aggregation of information leading to a higher statistical power and a more robust point estimate than that is possible from the measure derived from any individual study. Therefore, we have selected 13 independent studies to estimate *R*_0_ by meta-analysis in the paper. In these selected studies, these studies collected a random sample from a larger population with a COVID-19 disease. These random samples included different patient population with a different city, different countries, or different time periods, and so we proposed to use the random-effect meta-analysis model, which were developed by the papers **Hedges1985** and **DerSimonian1986**. The random-effects meta-analysis model is specified as

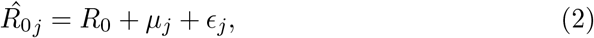

where *µ*_*j*_ *∼N* (0, *τ* ^2^) and 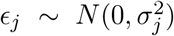 The parameter *τ* ^2^ represents the between-study variability and is often referred to as the heterogeneity parameter. It estimates the variability among the studies, beyond the sampling variability. Therefore, the target of inference is 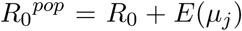. We use the weighted average as the estimator for 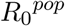 as follows:

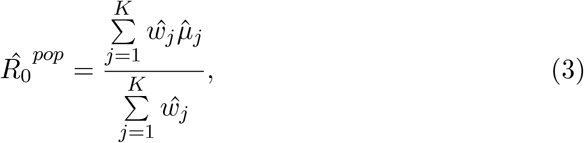

where 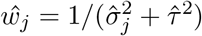

### 2.2 The infection rate per million population

COVID-19 is now a global pandemic, but some countries have been hit harder than others. The US now leads the world in overall cases. The government in each country has implemented its own policy requiring varying levels of isolation in their efforts to prevent COVID-19 spread. To compare the effectiveness of isolation policies across the countries in this section, we standardized the number of infectious cases by population size by (*Y*_*t*_*/P* × 100, 0000). The effectiveness of the isolation policy is negatively related to the effective reproductive number of *R*_*t*_. Therefore, the change curve of the standardized infectious rate over time can indicate the effectiveness of the isolation policy executed by the government.

By standardizing the reproduction rate to per million population, we can more feasibly compare the effect of the government closure policy across countries since it allows the reader to see both the number of the total population and the number of cases in a single measure. Additionally, since the health system capacity is proportional to the total population, the reproduction rate per million population is more likely to predict whether the health capacity in a specific country has been exceeded, which is important to know because this can lead to an unnecessary increase in morbidity and mortality. The statistical results will be given in result Section 2.2.

### 2.3 Healthcare capacity

It is possible to reduce the density of social contact, limited population isolation, and avoid the occurrence of super-transmission environment, the majority of single-door hospitals, as long as the public reduce the concentration of largescale activities, the peak of the disease to implement home-based office, students at home internet classes (online teaching) measures, can greatly reduce the spread of the disease. So the peak of the infection rate can be delayed or reduced after the intervention as shown in Figure 1, which is from two simulation data set with the same number of 10,000 infectious patients.

**Figure 1:**
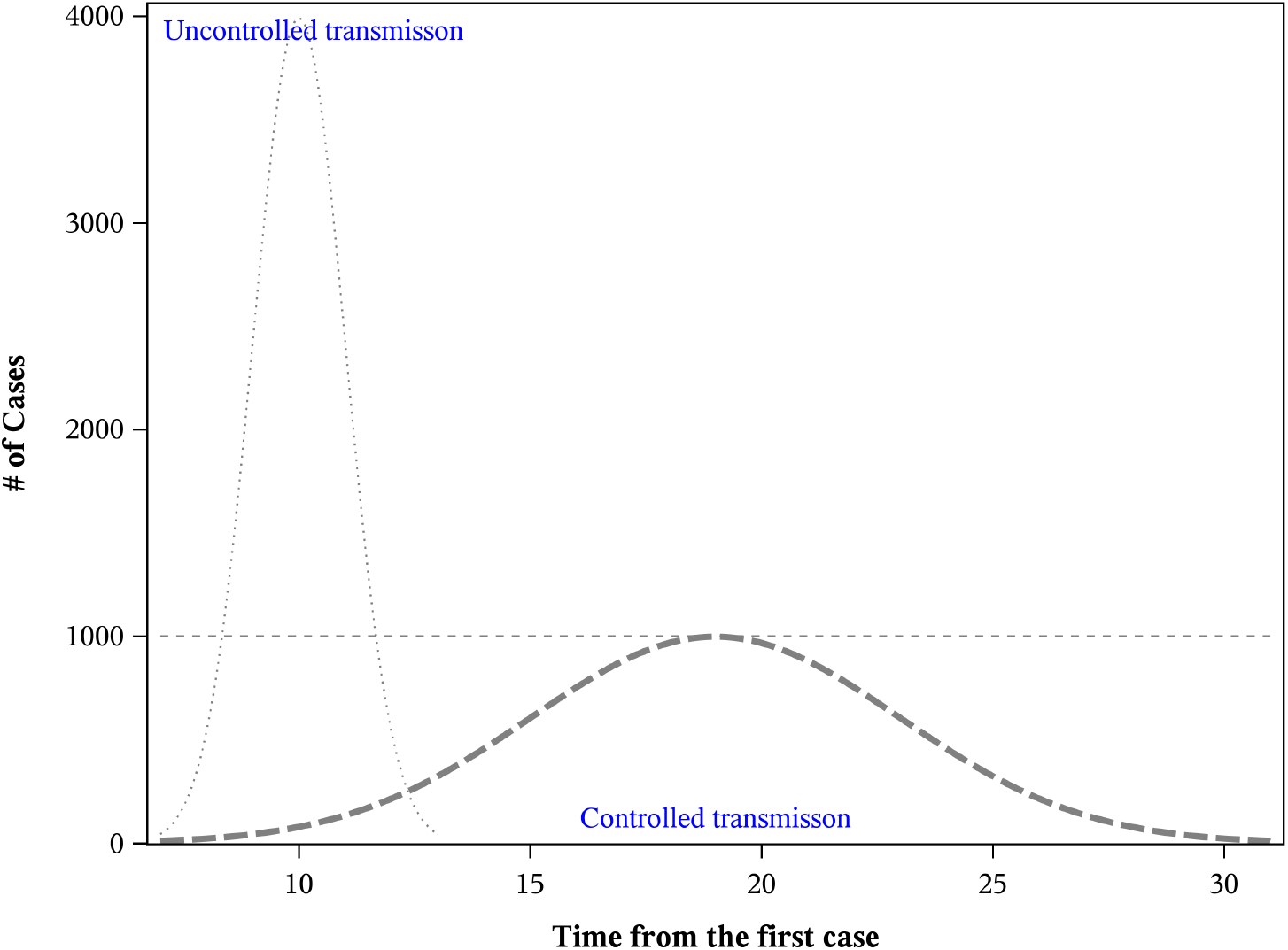
The flattered curve give how the infectious rate of COVID-19 can reduce the burdens of the health care system and the needed hospital capacity compared with the steep curve.

Although COVID-19 is likely to infect a large proportion of each country’s population, early and aggressive isolation policies should be effective in “flattening the curve” for the infection rate as shown in Figure 1. The “social distancing” measures have been put in place with the hope of delaying and reducing the overall peak of the infection rate curve. While the integral patient number of the normal curves is the same for both curves, the “flattened” curve never exceeds the capacity of the healthcare system, so that there are adequate resources to care for those with serious infections as well as those already requiring hospital care for other health conditions. For example, the Chinese government has made every effort to ensure that its citizens are not infected at the expense of the economy. In contrary to China, Europe, and the United States is relatively open at the beginning. We will give these comparisons in Section 3.3.

## 3 Results

This section applied the above-proposed methods to solve the research questions from the real COVID-19 data. We will show how the infectious number change with the effective reproductive because of the success of the isolation policy.

### 3.1 The basic reproduction number

Our estimate of *R*_0_ from meta-analysis is 3.05 as shown in Figure 2. Our value is larger than the range from 1.4 to 2.5 from WHO. We find that the stochastic and statistical methods for deriving *R*_0_ provide estimates that are reasonably comparable such as in the paper **Zhao2019**. Most mathematical methods produce the larger estimates, although the values from some of the mathematical methods were within the range produced the statistical and stochastic estimates. We find that the reproductive number of *R*_0_ of COVID-19 is larger than that of SARS.

**Figure 2:**
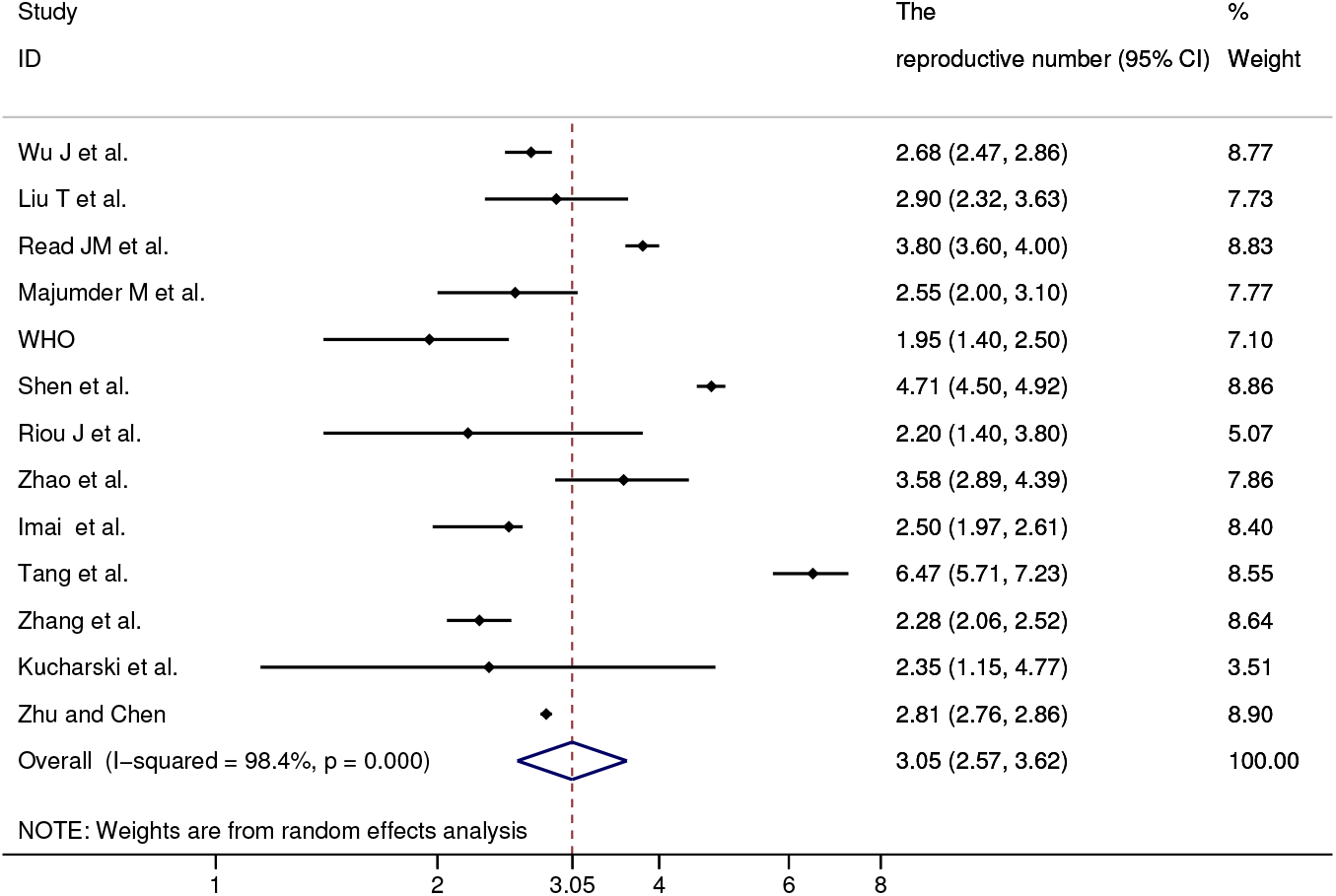
The reproduction number *R*_0_ from meta-analysis, which included 13 studies with the range between 1.95 and 6.47. Our robust estimate of *R*_0_ is 3.05, which is larger than 1.95 from WHO.

### 3.2 Infection rates per million population

This information graphic takes the ten most-affected countries in terms of total cases and calculates the cumulative infectious rate per million population. Using this measure, Italy has the most severe rate with 1,000 per million population as in Figure 3. The USA is increasing very fast and the pattern of its curve is similar to Italy. The curves of China and South Korean are very flat, which indicates that these two countries have controlled the epidemic in an effective way. Compared with South Korean, China has a relatively low infectious rate per million population due to its massive population and apparent containment for the time being.

**Figure 3:**
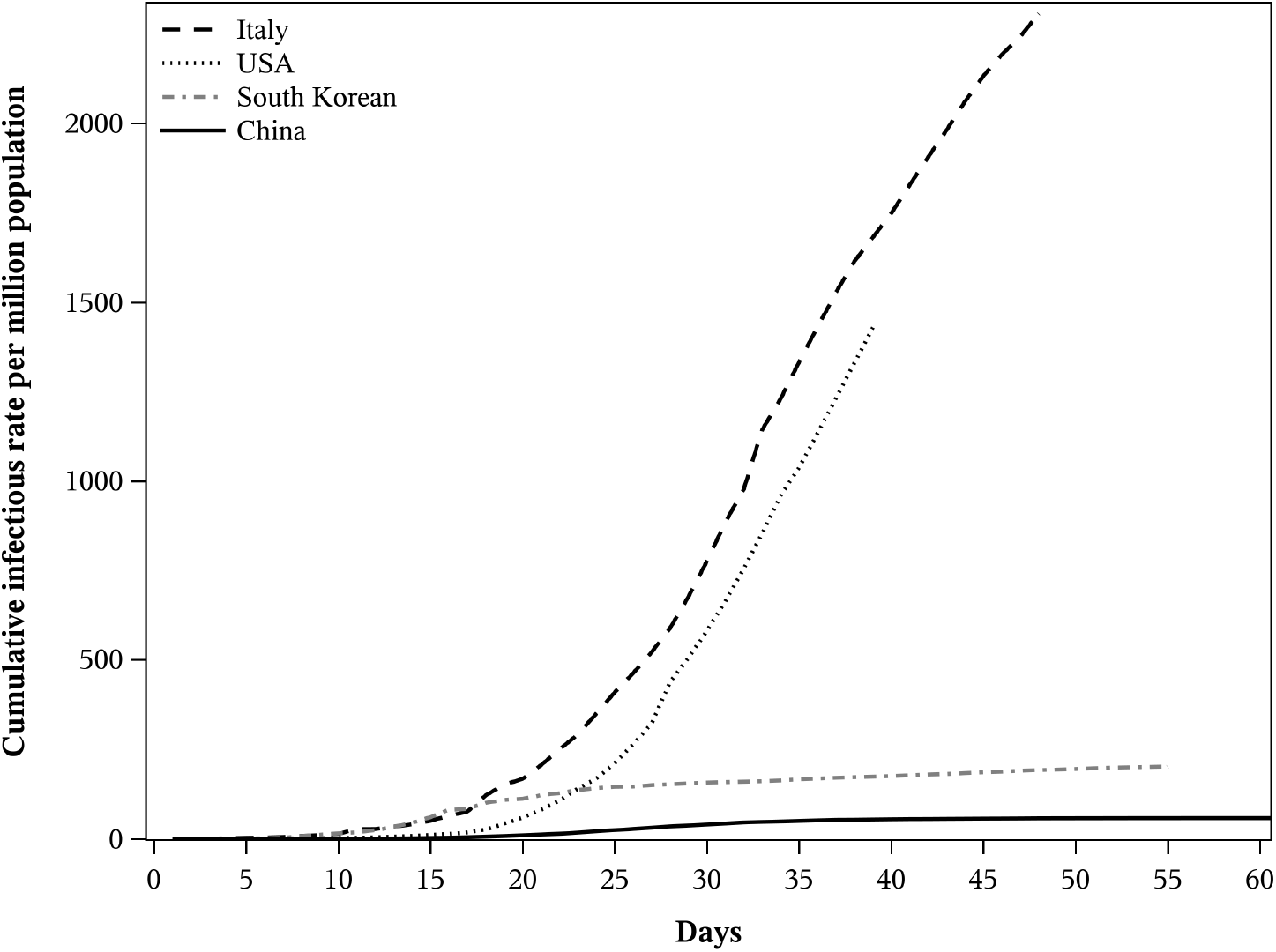
The infection rate curves per million population. The curves of China and South Korean were becoming very flat after 29 days, which indicates that both countries have effectively controlled the COVID-19 spreading. The curves of Italy and USA are at a similar climbing pattern.

Compared with the cumulative infectious rate, the incidence rate per million is a measure of the number of new cases per unit of time in the population. This paper also gives a comparison of the effectiveness of the isolation policy over time between China and the USA from the daily incidence infectious rate per million population over time. The epidemic process in China has been controlled in an effective way as shown in the upper panel in Figure 4. The solid curve in Figure 4 is the real infectious incidence rate from the real China data. The effective reproduction number decays in the dashed line, which is assumed as the exponential distribution with the basic reproduction number *R*_0_ (3.05) from meta-analysis at the start point, then reach 1:00 at the peak point. The decay curve indicates that the isolation policy in China was very successful since the effective reproduction number has dropped to 1:00 at peak time.

**Figure 4:**
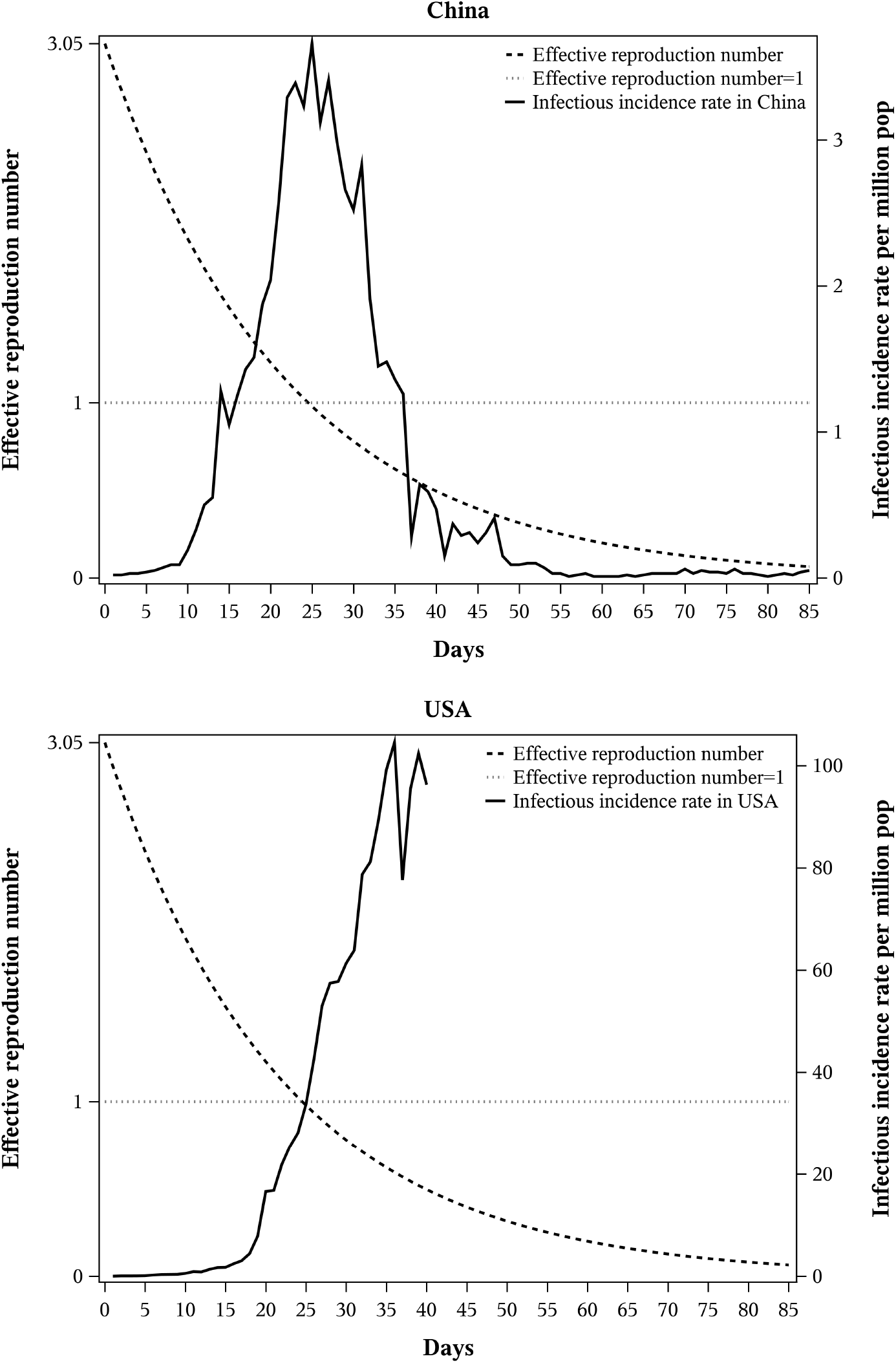
The effective reproduction number and the daily infectious number in China and the USA. The predicted dash line is the decay curvy of the effective reproductive number from China’s real data. The dotted line is a reference line when the effective reproductive number arrives one. The solid line in the above panel is the infectious number in China, and the solid line in the lower panel is the infectious number in the USA. The peak day was at about 26 days after Dec 20 in China while the USA is still rising after 26 days.

If the USA has similar effects of the isolation policy as China, then the effective reproduction number should decay as the dashed line as shown in the lower panel in Figure 4. However, the date to reach the peak of the number of the infectious number appears well beyond 26 days in the US. In fact, the US is still rising at an exponential rate, exposing an ineffective current isolation policy.

### 3.3 Healthcare capacity

As mentioned in Section 2.3, the peak of the infection rate can be delayed or reduced after the intervention such as the social distancing measures has been put in place. However, if the peak curve exceeds the capacity of the healthcare system without a successful intervention, especially for the heavy burden of serious infectious patients in the intensive care unit, then it causes a high mortality rate. This section wants to apply the pandemic theory to the real data of COVID-19 so that we can justify the fact why the mortality rate was low in China.

We give the daily infectious rate curve per million population against time in Figure 5. The dotted line is the daily infection rate per million population in China and the solid line is the daily infection rate per million population in Italy. Italy has reached 108.3 per million population compared with 3.5 per million population in China. So we can see that Italy has a more steep curve than China from the graph. The curve shows that there were no immediate effective preventive measures to stop the disease in the community in Italy compared with China. Given that hospital capacity per million population is similar to China, the epidemic in Italy was far beyond its maximum national health capability. It may partially explain why the mortality rate of 11.9%(13, 155*/*110, 574) in Italy was much higher than the mortality rate of 3.7%(3, 312*/*88, 554) in China as far April 1 2020. In other words, government measures such as city lockdown have not been put in place to reduce the overall peak of the infection rate curve, which has been mentioned in Figure 1 in Section 2.3.

**Figure 5:**
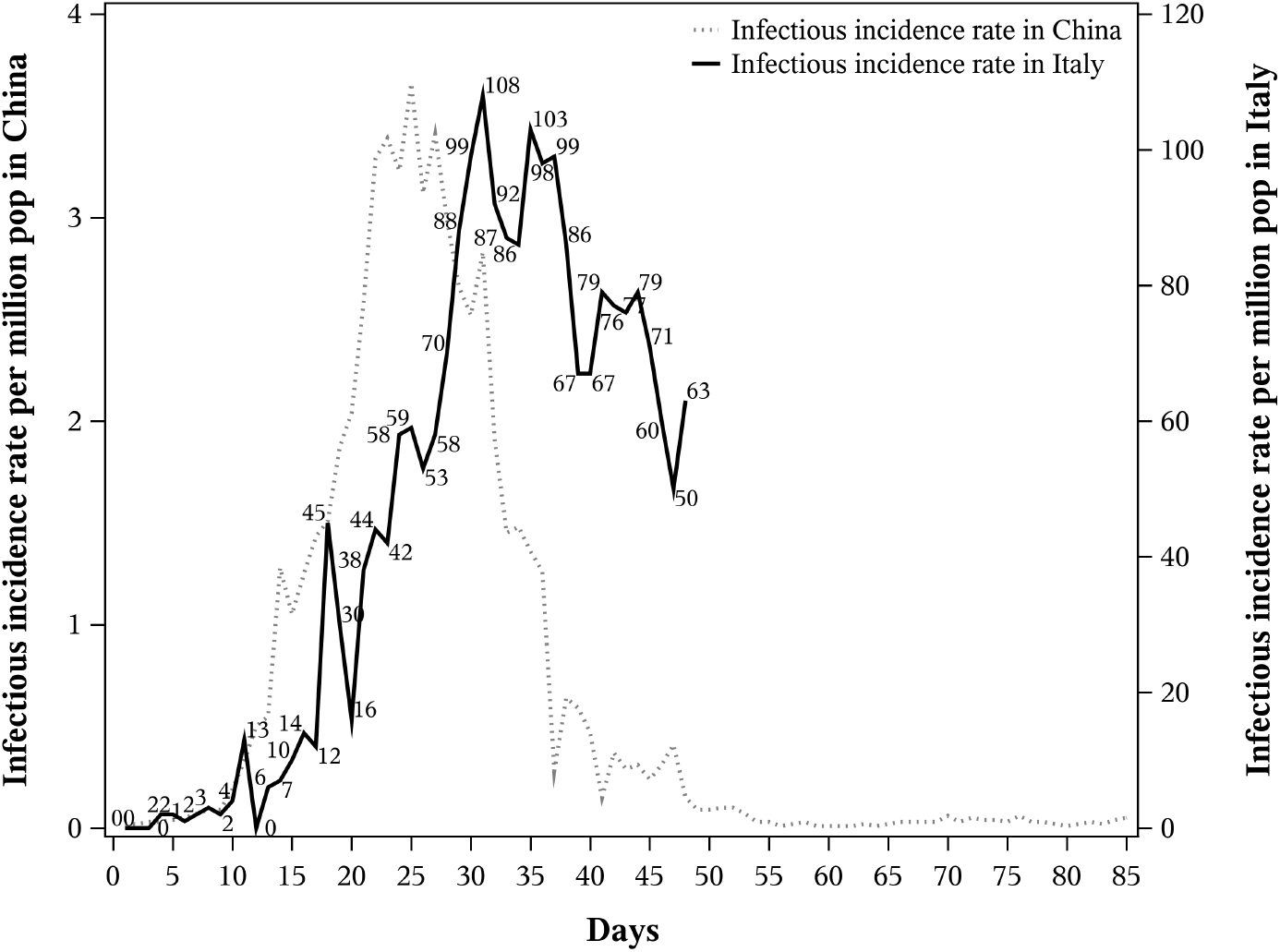
The dot line is the daily infection rate per million population in China and the solid line is the daily infection rate per million population in Italy. Italy has reached 108.3 per million population compared with 3.5 per million population in China

## 4 Conclusion

This paper developed statistical methods to solve some challenging problems in COVID-19 research. The estimate of the basic reproductive number has been carried out in many studies, which were involved in different samples in different cities and countries, different time periods for Wuhan infectious population, and simulation samples from statistical or mathematics models. So, the basic reproduction number varies from 1.95 to 6.47. According to the existing literature, we choose independent studies to estimate the *R*_0_ from a meta-analysis. we found that the estimated *R*_0_ for the COVID-19 from the meta-analysis is 3.05 (2.57, 3.62), which is considerably higher than the WHO estimate at 1.95. Also, we give the effective decay curve of *R*_*t*_ with an exponential distribution with the start point at 3.05 and at 1.00 at 26 days. The decay curve displays the whole process of COVID-19 in China with the successful isolation policy.

We find that the infection rate per million population is a good index to determine how the severity of COVID-19 in a specific country. Compared with the USA and Italy, the Chinese government is efficient and powerful to execute the isolation policy in this flight for COVID-19. There are several reasons for the Chinese successful isolation policy. Firstly, laboratory testing such as nucleic acid testing and CT scans have been widely applied in China, which was helpful for the identity and the isolation of COVID-19 patients at the right time and the right place. The timely screening of suspected COVID-19 patients can reduce the peak of the infectious rate of COVID-19, and then can reduce the burdens of the health care system in the whole country. Secondly, the large scale and severe exclosure policies were carried out to isolate infected people and quarantine their contact with healthy people. Finally, if the hospital system can increase the health capacity to take care of more patients by quickly building up mobile cabin hospitals, which can partially reduce the mortality rate.

The paper is applying the proposed methods to the COVID-19 research based on the available data and literature. The proposed model results such as *R*_0_ from the meta-analysis are driven by these existing data information and literature. As the outbreak is continually expanding to more regions in the world, the size of peak value and peak time may depend on a number of factors including the speed of diagnoses and hospitalization of confirmed cases. Therefore, updating the model results may need to be refined in the future.

## Data Availability

https://www.unmc.edu/publichealth/departments/biostatistics/facultyandstaff/jianghu-dong.html

